# Genomic map of blood group alleles in Malaysian indigenous Orang Asli population from whole genome sequences

**DOI:** 10.1101/2021.12.04.21267232

**Authors:** Mercy Rophina, Lay Kek Teh, Sridhar Sivasubbu, Vinod Scaria, Mohd Zaki Salleh

## Abstract

**Purpose:** Differences in the distribution of RBC antigens defining the blood group types among different populations have been well established. However, very few studies exist that have explored the blood group profiles of indigenous populations worldwide. With the rapid advent of next generation sequencing techniques and availability of population scale genomic datasets, we have successfully explored the blood group profiles of the Orang Aslis, who are the indigenous population of Malaysia and provide a systematic comparison of the same with major global population datasets.

**Methods:** Variant call files from whole genome sequence data (hg19) of 114 Orang Asli were retrieved from The Orang Asli Genome Project (OAGP). Systematic variant annotations were performed using ANNOVAR and only those variants spanning genes of 43 blood group systems and transcription factors KLF1 and GATA1 were filtered. Blood group associated allele and phenotype frequencies were determined and were duly compared with other datasets including Singapore Sequencing Malay Project (SSMP), aboriginal western desert Australians and global population datasets including The 1000 Genomes Project and gnomAD.

**Results:** This study reports 4 alleles *(rs12075, rs7683365, rs586178 and rs2298720) of* DUFFY, MNS, RH and KIDD blood group systems which were significantly distinct between indigenous Orang Asli and cosmopolitan Malaysians. Eighteen (18) alleles which belong to 14 blood group systems were found distinct in comparison to global population datasets. Although not much significant differences were observed in phenotypes of most blood group systems, major insights were observed on comparing Orang Asli with aboriginal Australians and cosmopolitan Malaysians.

**Conclusion:** This study serves as the first of its kind to utilize genomic data to interpret blood group antigen profiles of the Orang Asli population. In addition, systematic comparison of blood group profiles with related populations were also analysed and documented.

## Introduction

There are over 43 blood group systems in the world, and differences in the distribution of blood antigens and blood groups between populations have been well established. The antigenic determinants expressed on the surface of the Red Blood Cells (RBCs) are often regulated either by a single gene or by closely linked homologous genes. 345 unique human blood group antigens defined by about 1700 alleles across 50 genes have been recognized and duly approved by the International Society for Blood Transfusion (ISBT) till date.^1^ This diversity has enormous implications especially in countries which have diverse populations since. Mismatches in blood group antigens can lead to clinically significant alloimmunization and adversities during transfusion or pregnancy^2,3^. Accurate and extensive characterization of blood group antigen profiles to ensure safe and effective blood transfusions therefore becomes important. Owing to the rapid discovery of novel RBC antigens and their underlying genetic diversities, conventional serological techniques and medium throughput DNA assays become inadequate. Utilization of Next generation sequencing (NGS) based whole genome or exome data to extensively evaluate human RBC antigens encoding genes has been explored in recent years. ^4,5,6^

Malaysia is an abode of diverse populations in Southeast Asia, comprising 3 major ethnic groups of the Malays, Chinese and Indians alongside minority groups of the aboriginal Orang Asli and natives in the East of Malaysia. These native populations have remained underrepresented in major global population genome sequencing projects. Recent years have witnessed the efforts of understanding the genetic architecture of these native populations using high throughput DNA sequencing techniques.^7,8,9,10^ The Orang Asli population representing ∼0.7% of peninsular Malaysia comprises three major tribes namely Negrito, Senoi and Proto-Malays. Each of the tribes are further divided into subtribes based mainly on linguistic, physical, economical and cultural differences. The subtribes of Negrito were found historically associated with the initial wave of modern humans who migrated out of Africa ∼25,000 to ∼60,000 years ago forming the earliest descendants of Peninsular Malaysia.^11,12,13^ Genome sequencing data of these indigenous groups were generated by The Orang Asli Genome Project (OAGP) which was initiated to unravel the genomic architecture and environmental impacts in selection pressures. Although there have been a handful of efforts in deciphering various genetic signatures of this semi-isolated population, little is known regarding the prevailing blood group profiles.

A recent study by Schoeman and colleagues portraying distinct blood group profiles of indigenous western desert Australians^14^ has provided a systematic way of assessing genomic data to elucidate the distribution of blood group antigens in various indigenous populations. In this study, we aim to curate and annotate the comprehensive collection of blood alleles prevailing in the aboriginal Orang Asli population along with systematic prediction of complete blood group phenotypes from whole genome data. In addition, we also intend to filter population specific novel and rare variants with potential impacts in blood group profiles.

## Materials and Methods

### Reference datasets of human blood group genes and alleles

Genomic coordinates (GRCh37/hg19) of 50 genes associated with 43 human blood groups and 2 erythroid specific transcription factors were duly fetched from Locus Genomic Reference.^15^ Detailed summary of genomic coordinates is tabulated in **Table 1**. A systematically compiled reference data comprising ISBT approved blood group related alleles were fetched and documented in a pre-formatted template.^16^ The reference dataset extensively includes Single Nucleotide Variations (SNVs), Insertions, Deletions, Copy Number Variations (CNVs) and combination mutations.

**Table 1.**
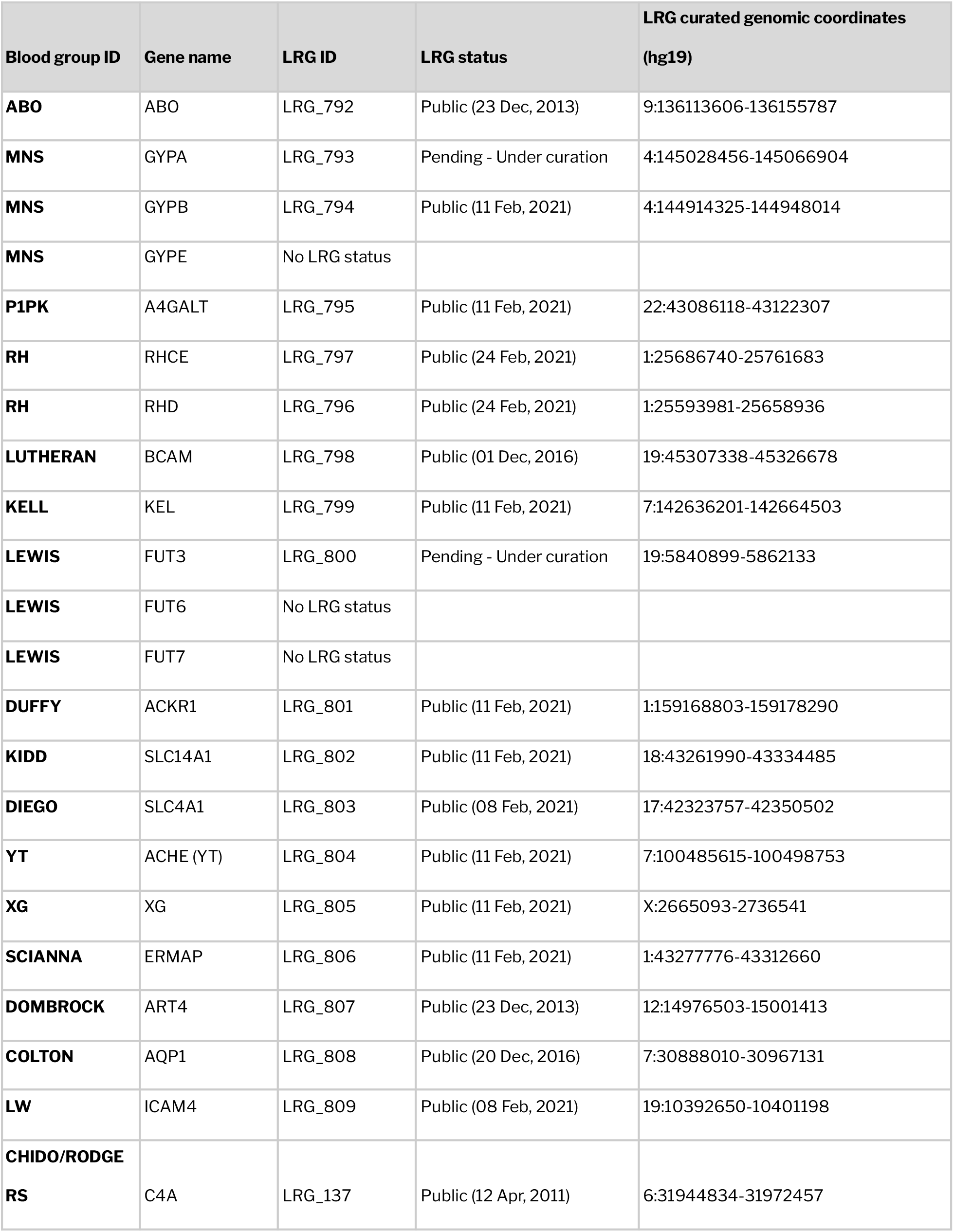
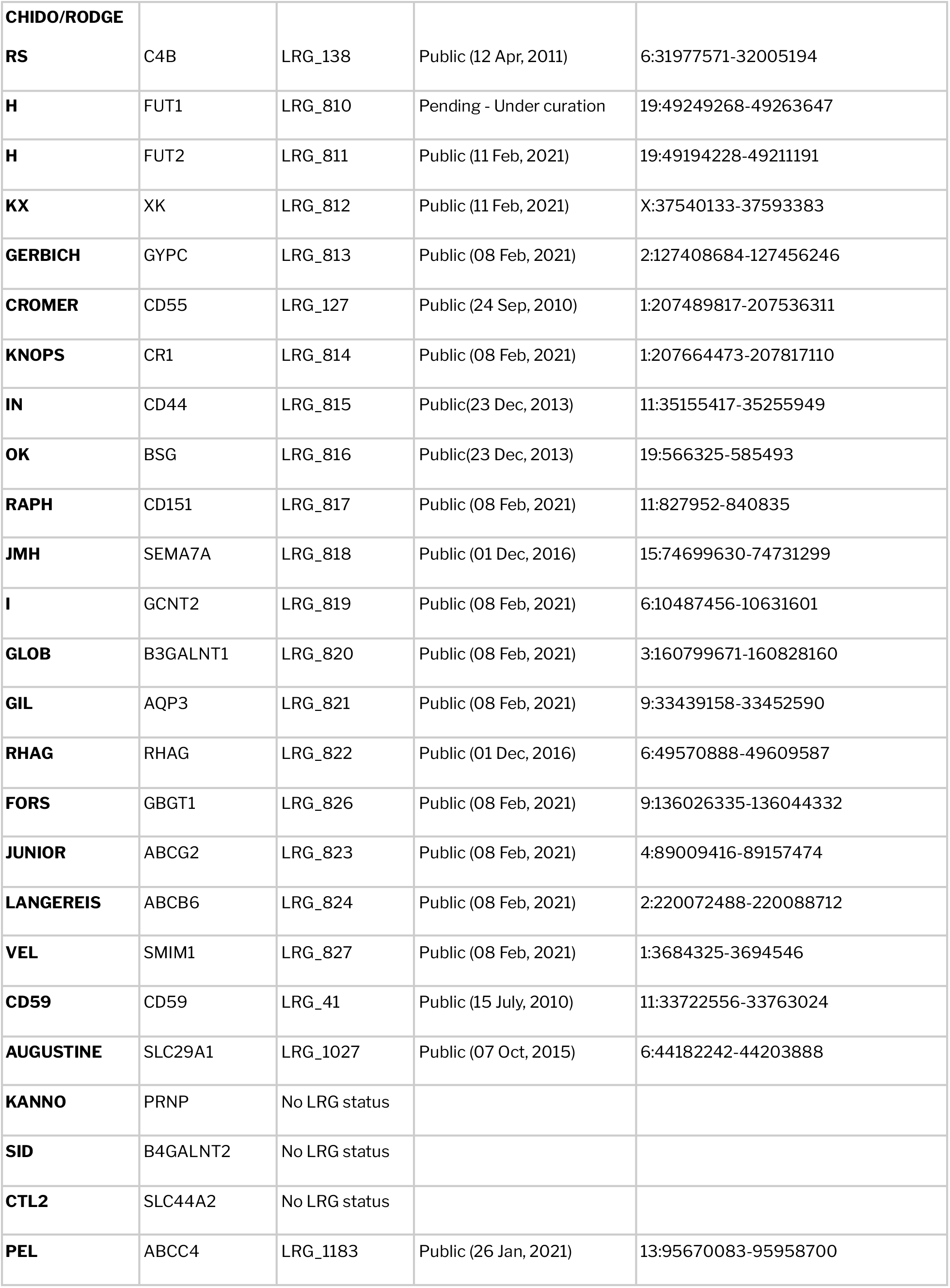
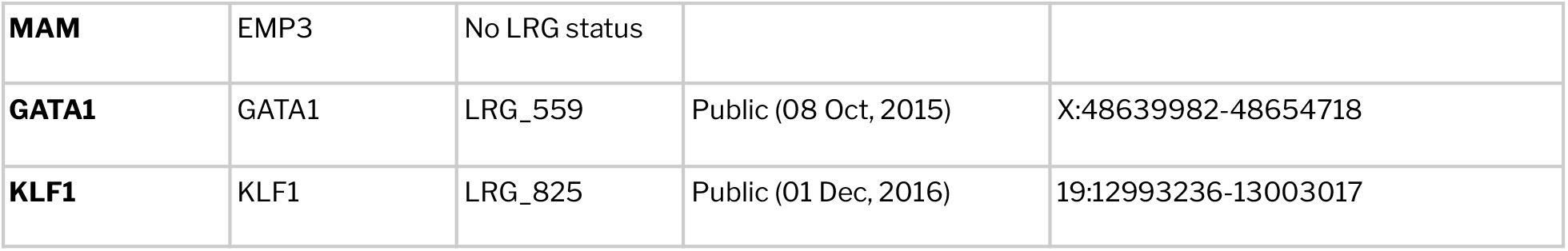
Summary of LRG genomic coordinates (hg19) for all the human blood group associated genes

### Genomic variation datasets

Genome sequencing data generated by The Orang Asli Genome Project (OAGP), was used in the study. The dataset comprised of sequence variations of 114 Malaysian Orang Asli individuals including the major subtribes namely Negrito *(Bateq - 23, Lanoh - 16, Kensiu - 20)*, Senoi *(Che Wong - 19, Semai - 17)* and Proto Malay *(Kanaq - 19)*. OAGP comprises a total of 21089667 unique genetic variations. In addition, genome sequencing data of 96 healthy Malays contributing to 13539289 unique variants were obtained from Singapore Sequencing Malay Project (SSMP) ^17^ and were used for comparison in the study. All the datasets corresponded to Human genome 19 (GRCh37/hg19) assembly. Comprehensive list of genetic variations was retrieved from the variant call format (VCF) files and were used for further analyses.

### Data processing and annotation

The initial level of data processing involved fetching all variants which were found to span the LRG coordinates of human blood group related genes and erythroid specific transcription factors. All filtered variants were annotated for their functional consequences using Annotate Variation (ANNOVAR).^18^ An extensive range of computational tools including SIFT,^19^ Polyphen,^20^ LRT, MutationTaster, Mutation Assessor,^21^ FATHMM,^22^ PROVEAN,^23^ CADD,^24^ GERP,^25^ PhyloP ^26^ and PhastCons were used to assess the functional impact of the variations.

### Identification of known and novel blood group alleles

The second level of data analysis was aimed at classifying the filtered variants into those with known blood group phenotype associations and novel/rare variants. Blood group phenotype associated variants were primarily retrieved from ISBT.^27^ A preformatted compilation of human blood group associated alleles was also obtained ^16^ and used for comparison. Phenotypes of blood group systems with no reported blood group related variants were conferred the same nomenclature as that of the reference genome as described previously. ^28^

In addition, a list of potentially novel variants was fetched based on the SNP identification numbers (dbSNP ID). A variant was termed potentially novel if it lacked the dbSNP ID. Exonic novel variants were further filtered based on their functional impact predicted using a range of computational tools and minor allele frequencies. Variants with MAF < 5% with absence of reported blood phenotypes were deemed as rare variants. A schematic representation of the methodology followed is shown in **Figure 1**. Complete blood group profiles were also predicted for each sample used in the study.

**Figure 1.**
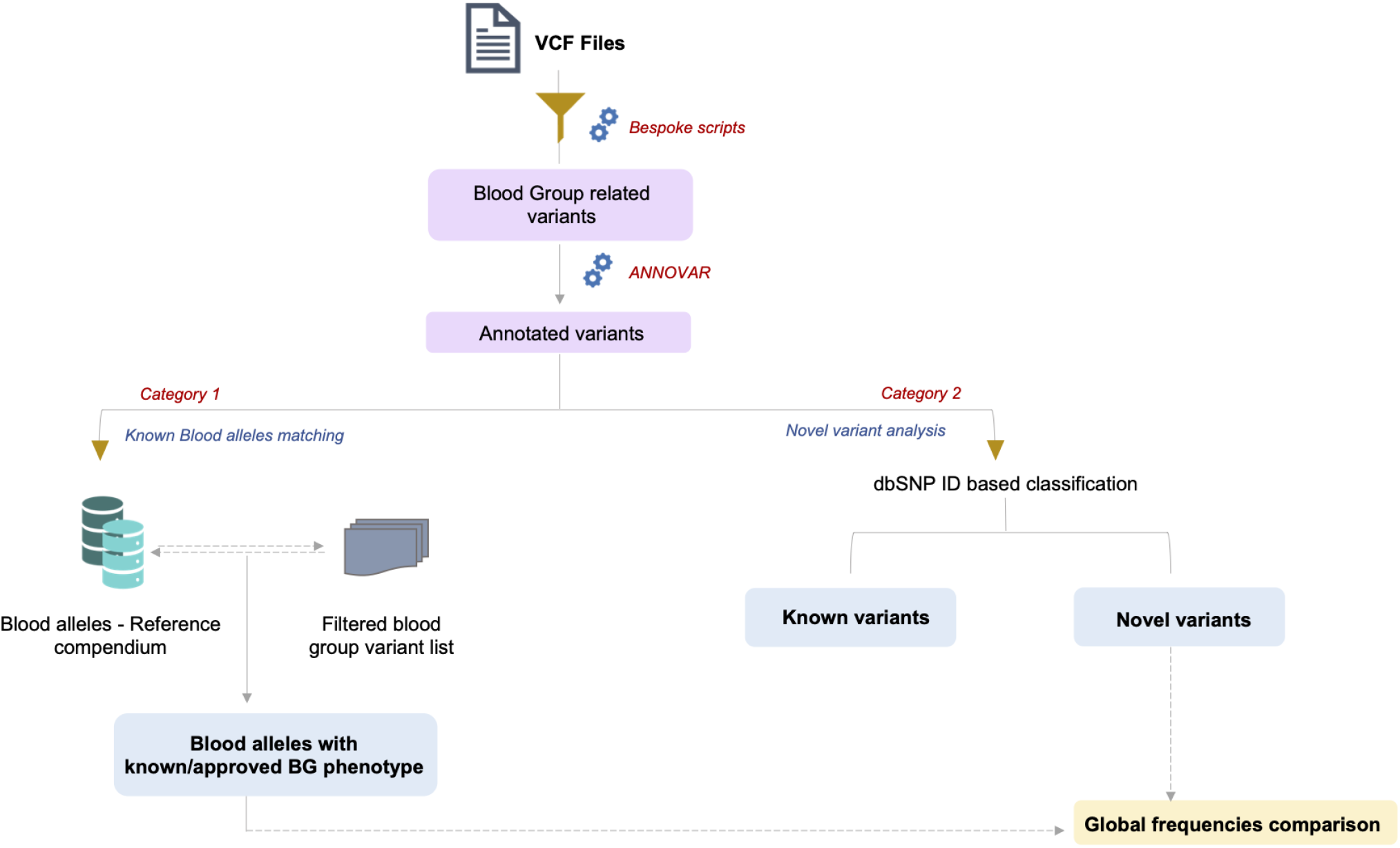
Schematic representation of the methodology followed in the study

### Estimation and comparison of allele frequencies

Filtered blood group associated variants from all the datasets were systematically compiled in Variant Call Format with corresponding genotype information. Allele frequencies were estimated using PLINK.^29^ Summary on the number of samples with homozygous and heterozygous genotypes were also generated using bespoke scripts. In addition, allele frequencies of the variants were fetched from major global population datasets including 1000 Genomes project^30^, Exome Aggregation Consortium (ExAC v.0.3)^31^ and Genome Aggregation Database (gnomAD)^32^ and were used for comparison. In addition to the global comparison, frequencies of blood group variants were systematically compared and analysed between the Singaporean Malays and among the subtribes of Orang Asli.

### Statistical analysis

With the aim of identifying significantly distinct blood group alleles specific for Orang Asli population, minor allele frequencies were compared with other global populations and statistical significance was observed using Fisher’s exact test with a P-value < 0.05. Distinct differences in blood alleles among Orang Asli subtribes were also checked. Alleles filtered as significantly distinct were further checked for their clinical relevance in transfusion procedures and in pregnancy settings.

## Results

### Overview of blood group associated variants and annotations in study dataset

In the primary level analysis, all variants spanning the LRG coordinates of blood group related genes were fetched from the datasets. A total of 12542 and 9616 variants were filtered as potential blood group associated variants from the Orang Asli and SSMP datasets, respectively. **Supplementary Table 1** provides the complete summary of variant counts for each blood group system for the above mentioned datasets. In the Orang Asli dataset, about 226 of the total variants were detected across the exonic and splicing sites. Of these, 119 were found to be nonsynonymous SNVs, 80 synonymous SNVs and 1 stopgain mutation. A schematic representation of the variant summary and functional classifications across the datasets is shown in **Figure 2**.

**Figure 2.**
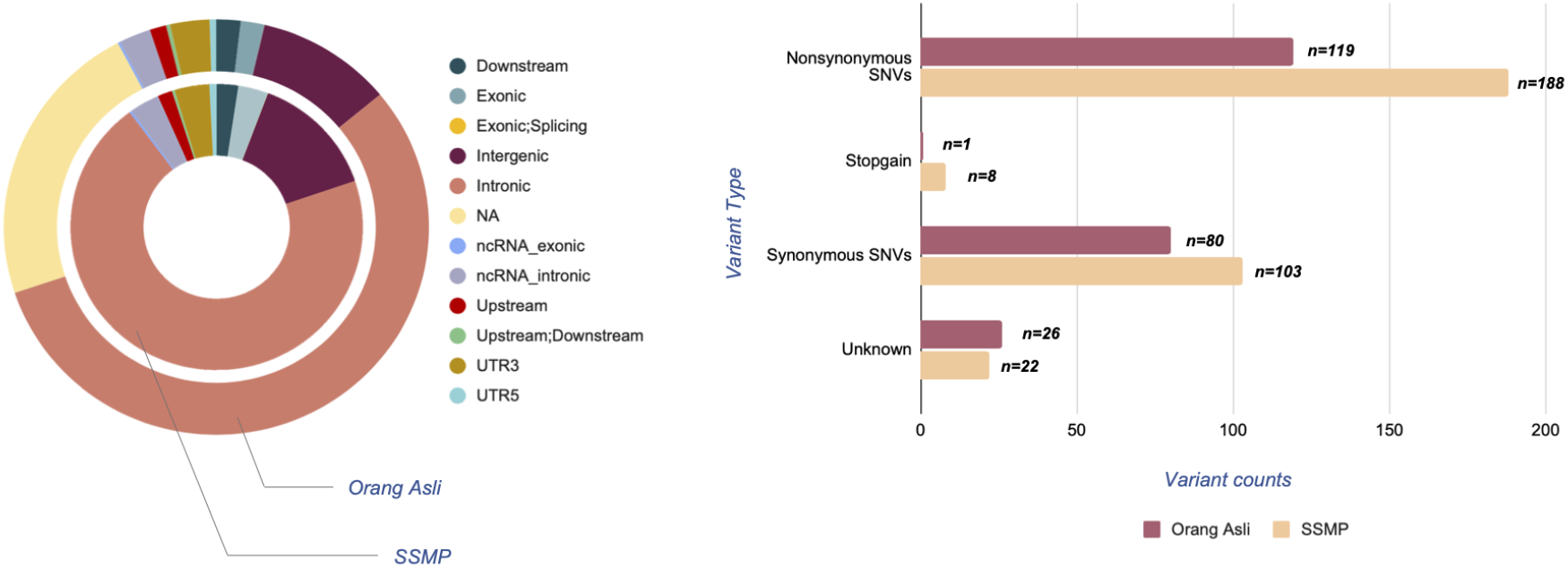
Complete overview of the blood group genes spanning variants and their corresponding functional classifications observed across the datasets used in the study

### Identification of blood group alleles and prediction of blood group phenotypes

Systematic comparison of filtered variants with reference resources revealed that a total of 33 variants belonging to 15 blood groups systems possessed blood group associated phenotypes. Twenty-one (21) of the total 33 variants were SNVs and the rest were combination mutations. For the remaining 28 blood groups including 2 erythroid specific transcription factors, the predicted phenotypes were reported the same as that of the human reference genome (hg19).^28^ Similar methodologies were followed in predicting blood group phenotypes of cosmopolitan Malaysian population dataset used in the study. **Table 2** details the phenotypes and genotypes of blood group systems reported to be the same as that of the reference genome in the Malaysian Orang Asli population. Comprehensive allele and phenotype frequencies of variants which were found to match reported blood group phenotypes are compiled in **Table 3. Supplementary Tables 2-4** provide the complete blood type profiles of each sample used in the study, a comprehensive compilation of each blood group system phenotype observed in the Orang Asli population and the distribution of blood phenotypes in Orang Asli subtribes, respectively.

**Table 2.** Tabulation of blood group phenotypes predicted to match the reference genome in 114 Malay Orang Asli samples.

| **ISBT Blood** |  |  |  |  |
| --- | --- | --- | --- | --- |
| **ISBT Blood Group** |  | **Group System** |  |  |
| **Blood Group System** | **ID** | **Number** | **Predicted Genotype call** | **Predicted Phenotype** |
| **DIEGO** | DI | 010 | DI*02/DI*02 | Di(a-b+) |
| **YT** | YT | 011 | YT*01/YT*01 | Yt(a+b-) |
| **DOMBROCK** | DO | 014 | DO*01/DO*01 | Do(a+b-) |
| **COLTON** | CO | 015 | CO*01.01/CO*01.01 | Co(a+b-) |
| **LW** | LW | 016 | LW*05/LW*05 | LW(a+b-) |
| **CHIDO/RODGERS** | CH/RG | 017 | C4B*03/C4B*03 | Ch+Rg- or CH:1,2,3,4,5,6<br>RG:-1,-2 |
| **GERBICH** | GE | 020 | GE*01/GE*01 | GE:2,3,4 |
| **CROMER** | CROM | 021 | CROM*01/CROM*01 | Cra+ |
| **IN** | IN | 023 | IN*02/IN*02 | In(a-b+) |
| **OK** | OK | 024 | OK*01.01/OK*01.01 | Ok(a+) |
| **RAPH** | RAPH | 025 | RAPH*01/RAPH*01 | MER2+ |
| **I** | I | 027 | GCNT2*01/GCNT2*01 | I |
| **GIL** | GIL | 029 | GIL*01/GIL*01 | GIL+ |
| **VEL** | VEL | 034 | VEL*01/VEL*01 | Vel+ |
| **CD59** | CD59 | 035 | CD59*01/CD59*01 | CD59.1+ |
| **AUGUSTINE** | AUG | 036 | AUG*01/AUG*01 | At(a+) |
| **KANNO** | KANNO | 037 | KANNO*01/KANNO*01 | KANNO1+ |
| **SID** | SID | 038 | SID*01/SID*01 | Sd(a+) |
| **CTL2** | CTL2 | 039 | CTL2*01/CTL2*01 | VER+ |
| **PEL** | PEL | 040 | ABCC4*01/ABCC4*01 | PEL+ |
| **MAM** | MAM | 041 | MAM*01/MAM*01 | MAM+ |
| **EMM** | PIGG | 042 | PIGG*01/PIGG*01 | Emm+ |
| **ABCC1** | ABCC1 | NA | NA | NA |
| **JMH** | SEMA7A | 026 | JMH*01 | JMH:1 or JMH+ |
| **RHAG** | RHAG | 030 | RHAG*01 | RHAG:1 or Duclos+ |
| **KLF1** | KLF1 | TF | KLF1*01 | Common |
| **XG** | XG | 012 | NA | NA |
| **KX** | XK | 019 | NA | NA |

**Table 3.**
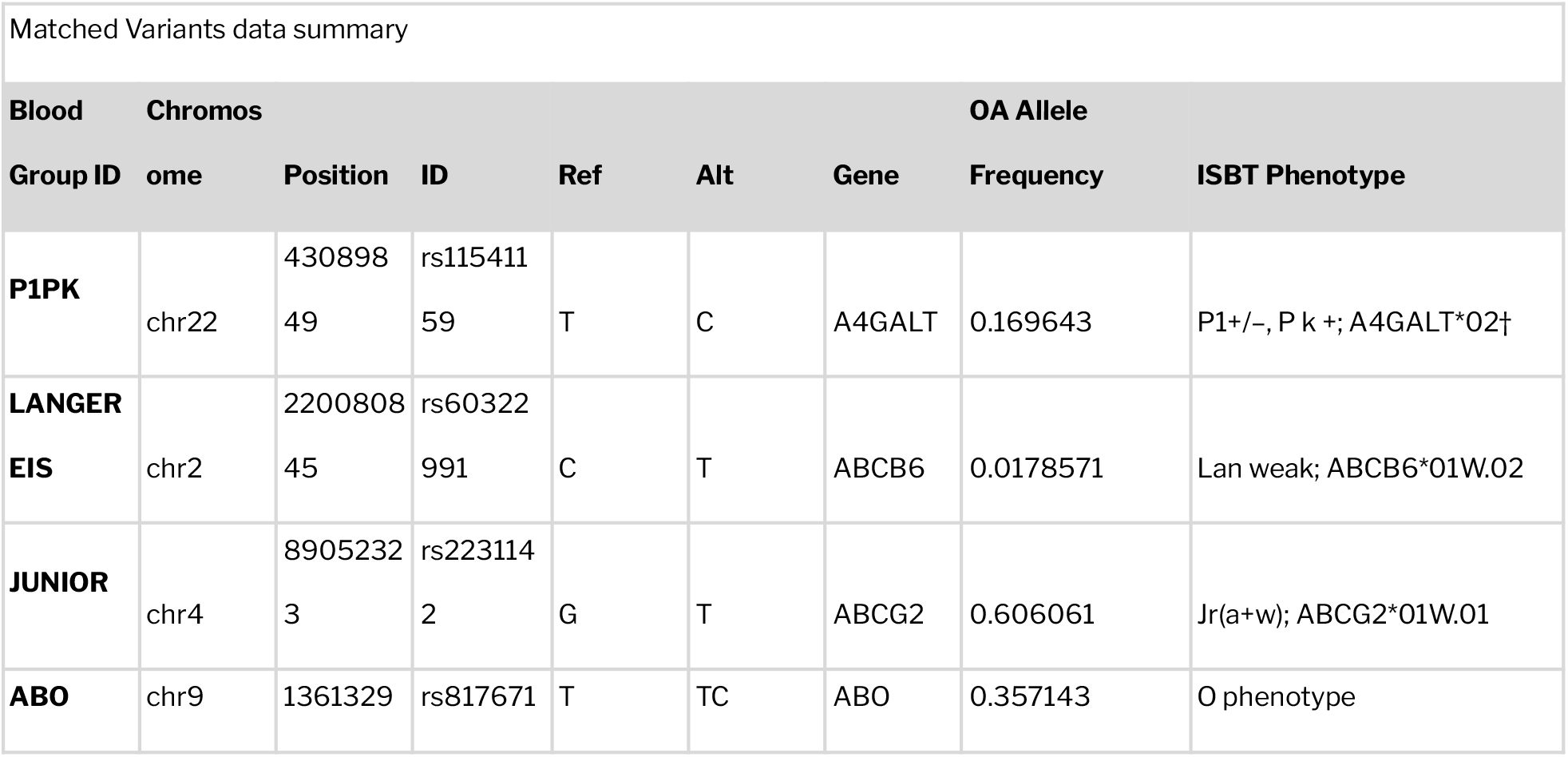
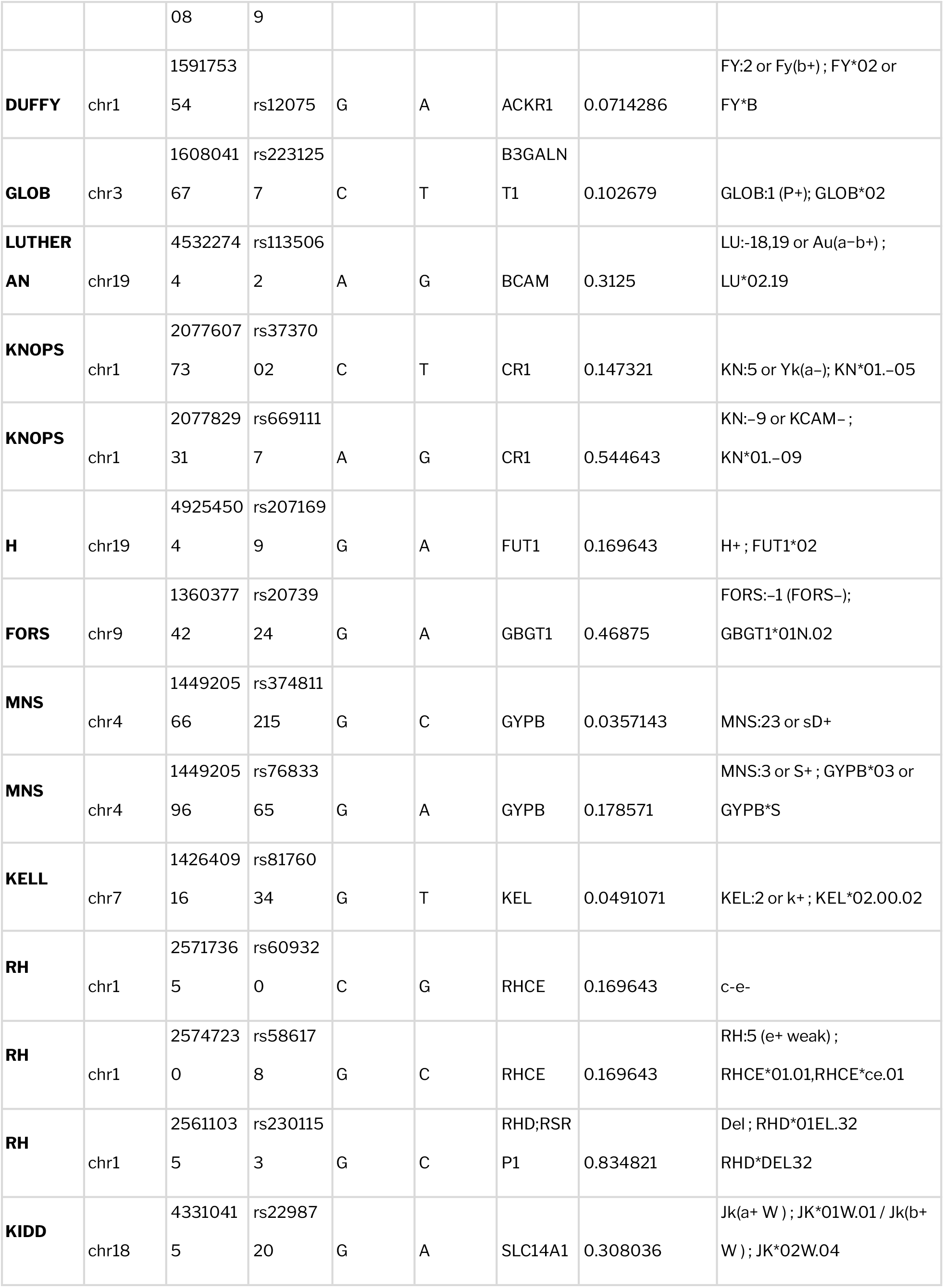
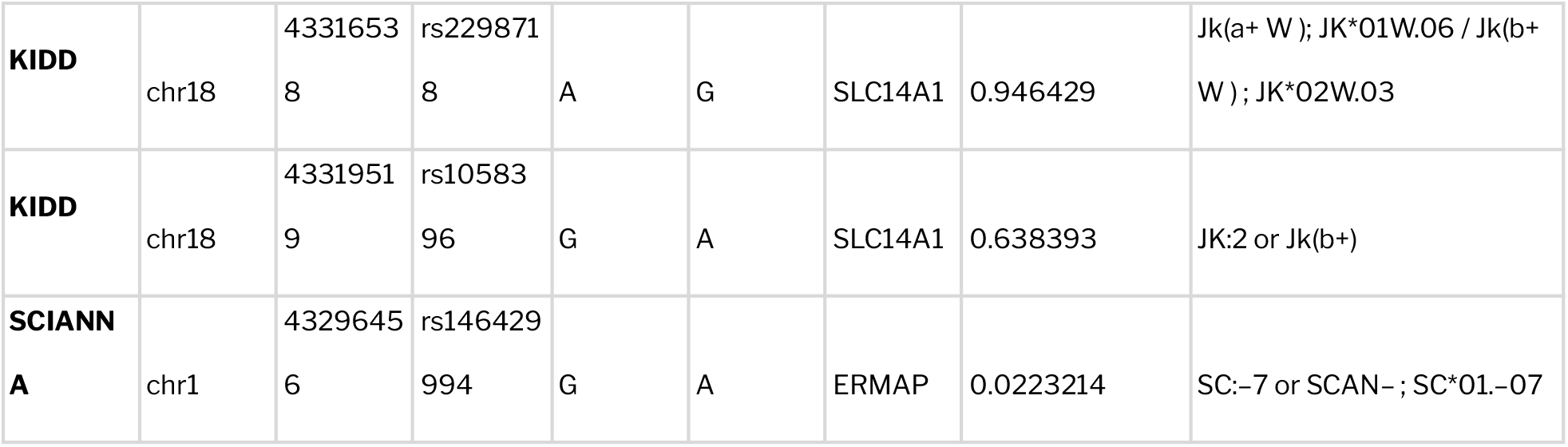
Tabulation of blood group alleles predicted to match ISBT approved phenotypes in 114 Malay Orang Asli samples.

### Impact of novel and rare variants in blood group profiles

Variant classification based on dbSNP identifiers revealed that a total of 3060 variants were found potentially novel. This systematically includes 21 exonic variants primarily belonging to LAN, PEL, JUNIOR, GLOB, LUTHERAN, CROMER, KNOPS, H, LEWIS, KELL, KLF1 and JMH blood groups with no global population frequencies reported and 6 variants of CROMER, KNOPS, JUNIOR, LEWIS and H blood groups which were computationally predicted to be deleterious by at least three or more tools. **Supplementary Figure 1** provides the distribution of novel variants filtered from various blood group systems. Description of the six novel variants predicted to have potential impacts on blood group profiles is provided in **Table 4**. There was a total of 74 rare blood group variants belonging to 25 unique blood group systems. Seventeen (17) variants out of the total were potentially novel which included 13 nonsynonymous SNVs and 4 synonymous SNVs. List of rare and potentially novel blood group variants are listed in **Supplementary Table 5**.

**Table 4.**
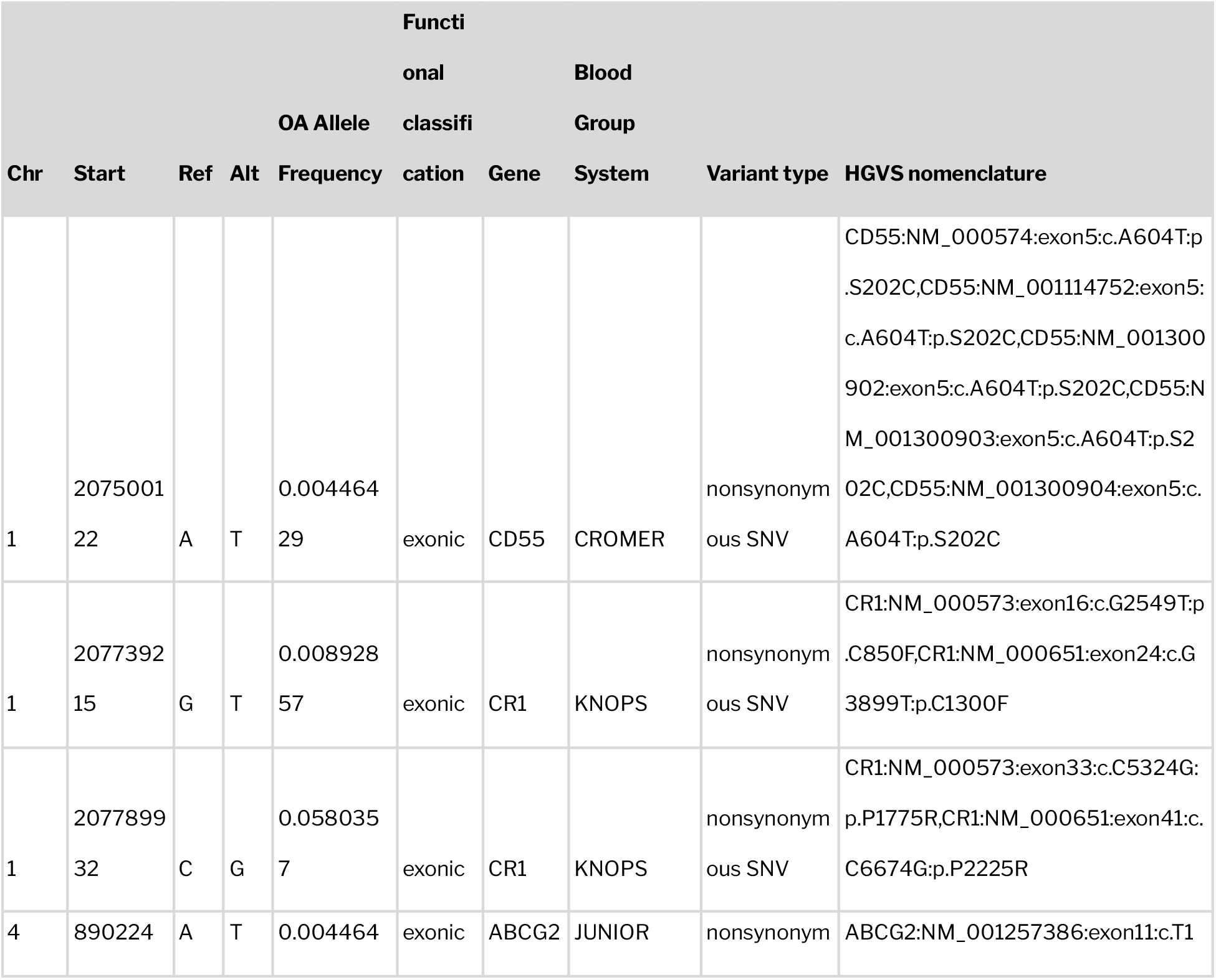
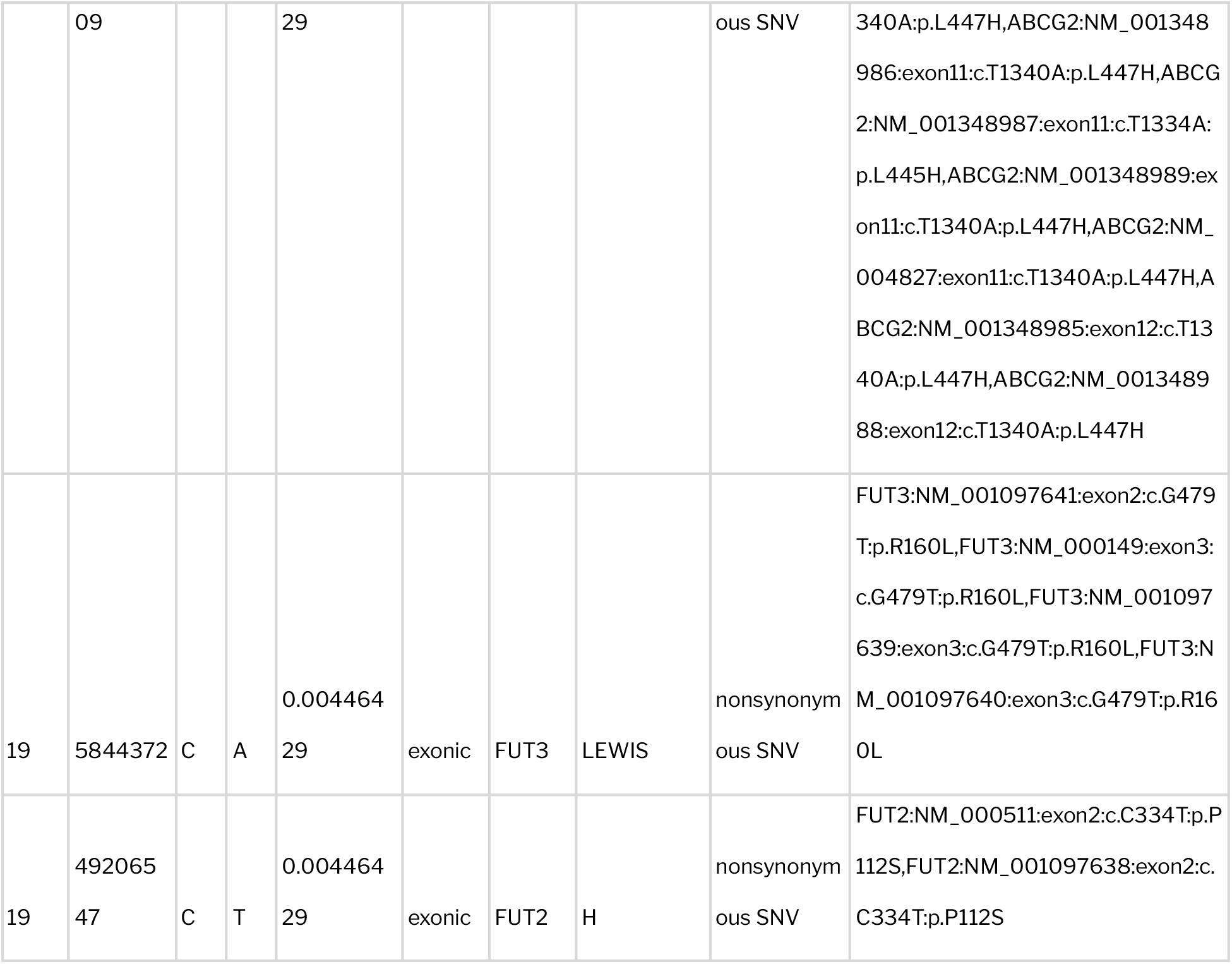
Description of 6 novel SNVs filtered in the dataset with predictions of potential impact in blood group profiles.

### Comparison of blood group profiles among sub-tribes and across datasets

Minor allele frequencies of blood group associated alleles were fetched from all datasets used in the study along with their corresponding frequencies in major global population datasets and significant differences across datasets were observed. **Supplementary Table 6** summarizes the frequencies of blood group alleles across various datasets. A total of 18 variants belonging to 14 blood groups were found significantly distinct in the Orang Asli population in comparison to global population datasets (1000 Genomes Project and gnomAD). List of these variants along with their P-value is tabulated in **Table 5**. In addition, 4 variants (rs12075, rs7683365, rs586178 and rs2298720) belonging to 4 unique blood group systems namely DUFFY, MNS, RH and KIDD were found distinctly different between the aboriginal Orang Aslis and cosmopolitan Malaysians. The observed P values corresponding to the variants are 0.0270, 0.0030, 0.0000 and 0.0162 respectively. A complete overview of the distinctly different blood group alleles and corresponding phenotypes is depicted in **Figures 3A and 3B**. Blood group allele frequencies distributed among the sub tribal populations of Orang Asli is shown in **Figure 3C**.

**Table 5.**
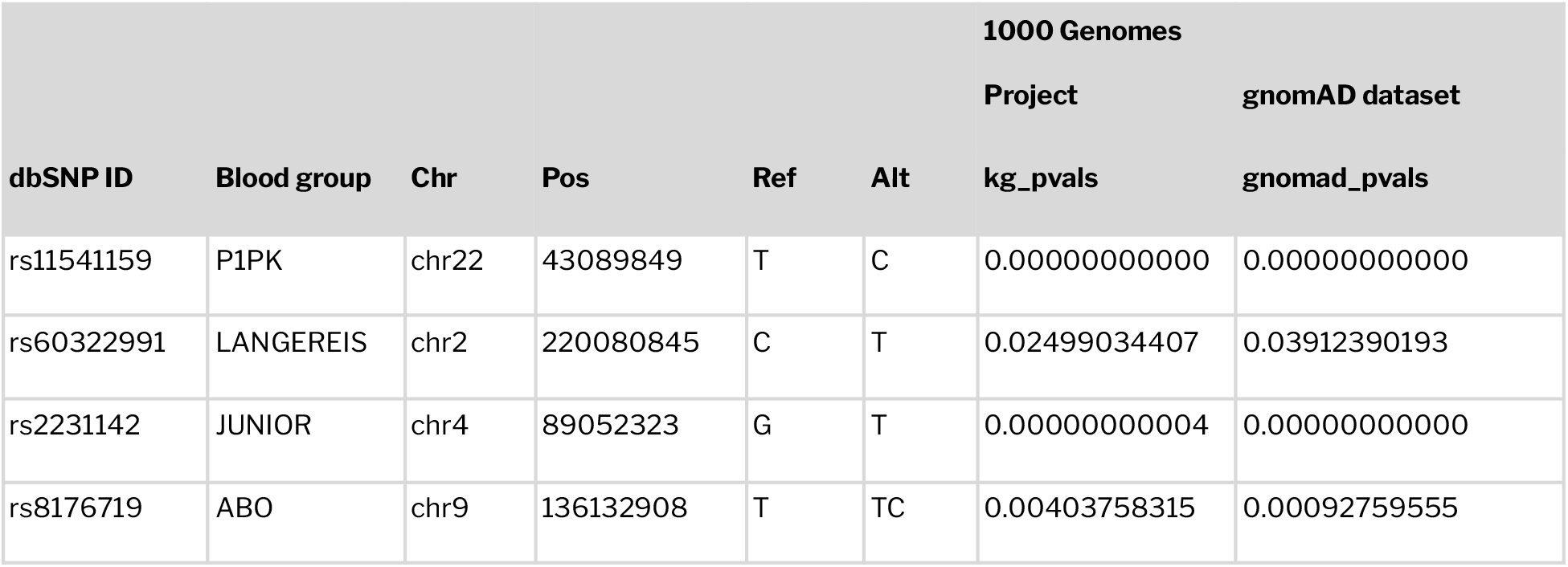
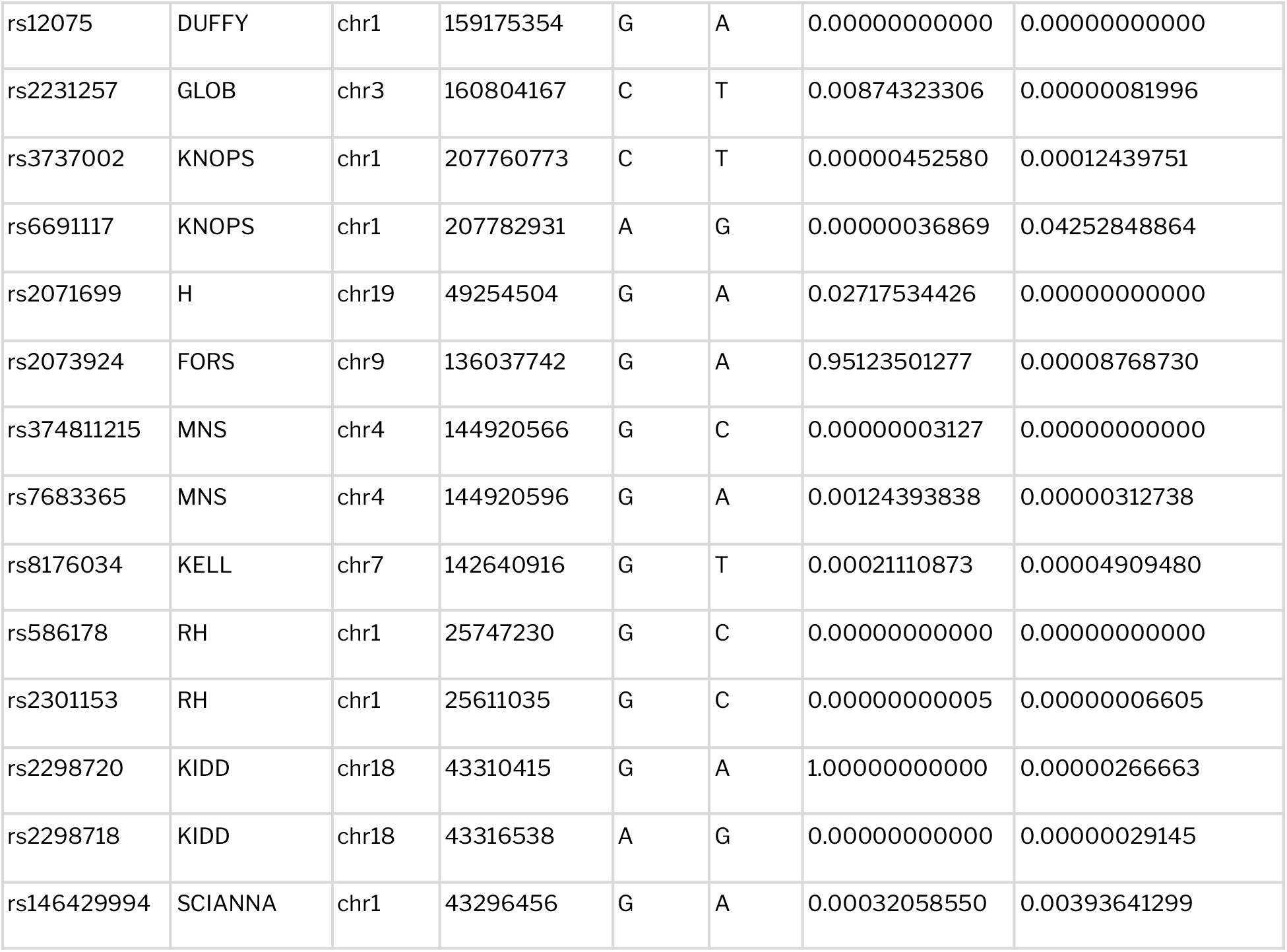
Summary of blood group alleles found significantly distinct between the aboriginal Malays and the global population datasets

**Figure 3.**
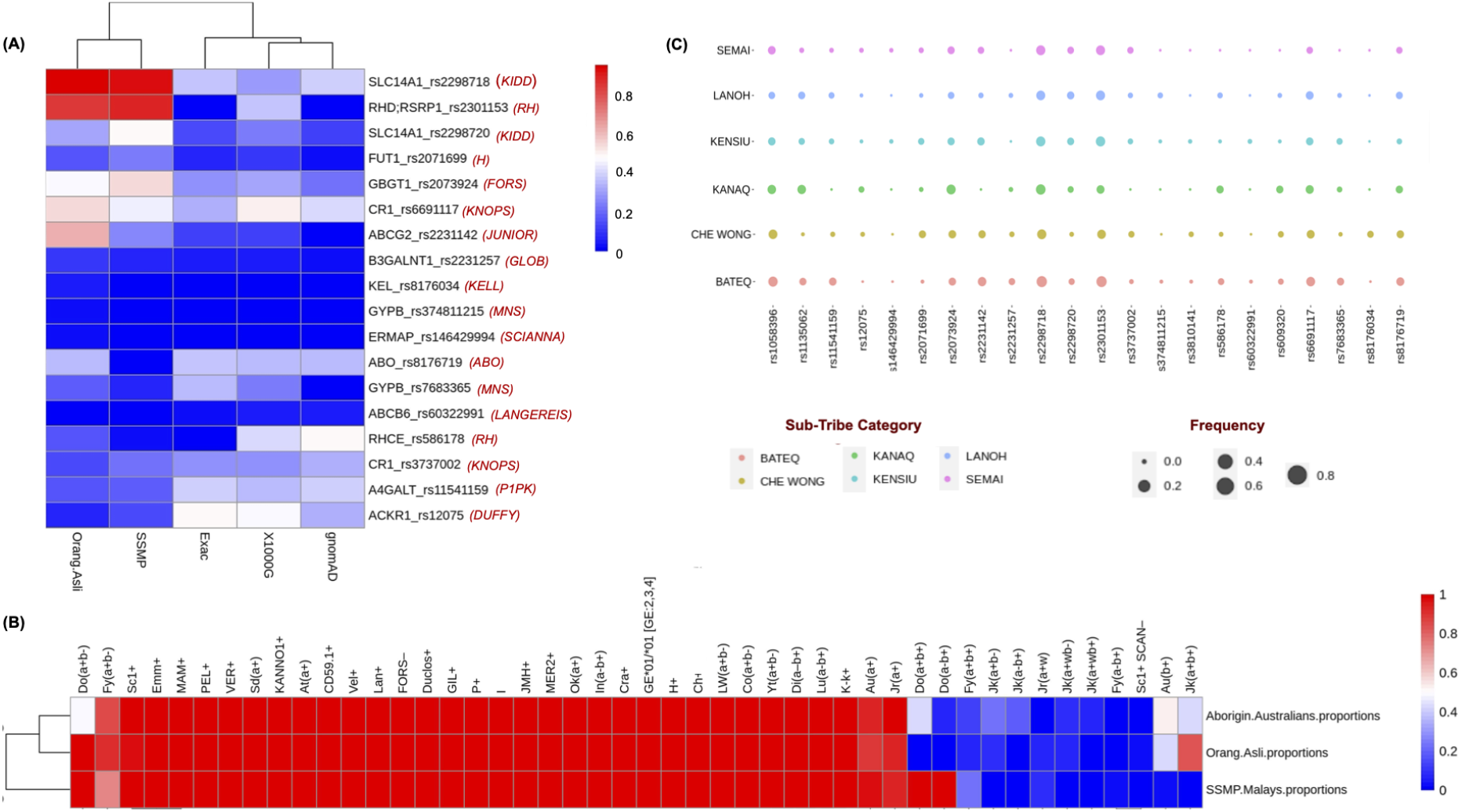
Distribution of blood group alleles and phenotypes among various global populations and among Orang Asli sub tribes. (A) Significantly distinct blood group alleles between Orang Aslis, cosmopolitan Malaysians and other global populations. (B) Pattern of distribution of blood group phenotypes among Orang Aslis, cosmopolitan Malaysians and aboroginal western desert Australians. © Distribution of blood group alleles in Orang Asli sub-tribal populations.

### Weak and partial antigens in Orang Aslis

There is a potential risk of hemolytic transfusion reactions in case of an antigen-negative recipient receiving an antigen-positive donor or the vice versa. Mistyping of weakly or partially expressed RBC antigens in donor RBCs as antigen-negative state can immensely alter the clinical phenotype thereby inducing adverse immune reactions. Interestingly in our study, we were able to observe weak alleles in Kidd and Junior blood group systems.

In Kidd blood group system, JK*01W.01 allele, which is predicted to weaken the antigen expression even in heterozygous genotype ^33^ and responsible for the JK^a+w^ phenotype, is observed in about 31% of the Orang Asli population. The observed allele frequency is found comparable to African *(1000 Genomes - 21%* ; *gnomAD genomes - 20%)*, East Asian *(1000 Genomes - 40%* ; *gnomAD genomes - 40%)* and South Asian *(1000 Genomes - 29%)* populations whereas distinctly varies from European *(1000 Genomes - 8%* ; *gnomAD genomes - 7%)* and American *(1000 Genomes - 19%* ; *gnomAD genomes - 18%)* populations.

Similarly, in Junior blood group system, the weak allele ABCG2*01W.01, manifesting the Jr^a+w^ phenotype,^34,35^ is observed in 36% of the indigenous population. Frequency of this allele was found to vary distinctly from rest of the global populations (*African* : *1000 Genomes - 1*.*3%, East Asian* : *1000 Genomes - 3*.*0%, South Asian* : *1000 Genomes - 9*.*7 %, European* : *1000 Genomes - 9*.*4%, American* : *1000 Genomes - 14%)*

## Discussion

This study serves first of its kind to provide the most comprehensive genetic blood group profiles of indigenous Malaysian Orang Asli population including all 43 human blood group systems and 2 erythroid specific transcription factors. Blood group alleles which are significantly different between the indigenous Orang Asli and Singaporean Malays as well as global populations were filtered. It was interesting to note that distribution of blood group allele frequencies was comparatively similar between the cosmopolitan Malaysians and Orang Asli than other global populations. In addition, although not many differences were observed in most blood group phenotypes, blood group systems with distinct changes in the manifested phenotypes among populations were also fetched. Evidence of similarities in blood group phenotypes between the Malaysian Orang Asli and the aboriginal western desert Australians were also observed. A precise compilation of alleles encoding weak/partial antigens, novel and rare alleles specific to Orang Asli was performed. Our analysis is mainly limited by the fact that RBC antigenic expressions regulated by large deletions and insertions (especially in RH and MNS blood groups) have not been profiled owing to the limitations in the datasets. In addition, the level of concordance with serology based phenotype predictions to investigate the novel and rare variants remains to be defined.

## Conclusion

Level and trend of healthcare settings often differs between indigenous and non-indigenous populations. Proportion of deaths avoidable through primary, secondary and tertiary services has always been observed higher in indigenous populations worldwide.^36^ There arises increased blood transfusion requirements in cases of chronic disorders. One of the early studies which was aimed at exploring the allelic diversity of human platelet antigens of this isolated population stated that obvious similarities and differences were observed between the Orang Asli and other major Malaysian subpopulations owing to their ancestral founders.^37^ This study emphasizes the importance of population scale sequencing efforts towards elucidating the comprehensive blood group antigen profiles of Malaysian Orang Asli population.

## Data Availability

All data produced in the present work are contained in the manuscript

## Author Contributions

VS conceived and designed the project with MZS and LKT. MR and VS contributed in writing the manuscript. All authors approved the final manuscript. Authors acknowledge funding from CSIR India and Ministry of Higher Education, Malaysia. The funders had no role in the preparation of the manuscript or decision to publish.

## Acknowlegments

The authors acknowledge Arvinden VR and Srashti Jyoti Agrawal for their constructive comments and suggestions.

## Supplementary Figures and Tables

**Supplementary Figure 1.**
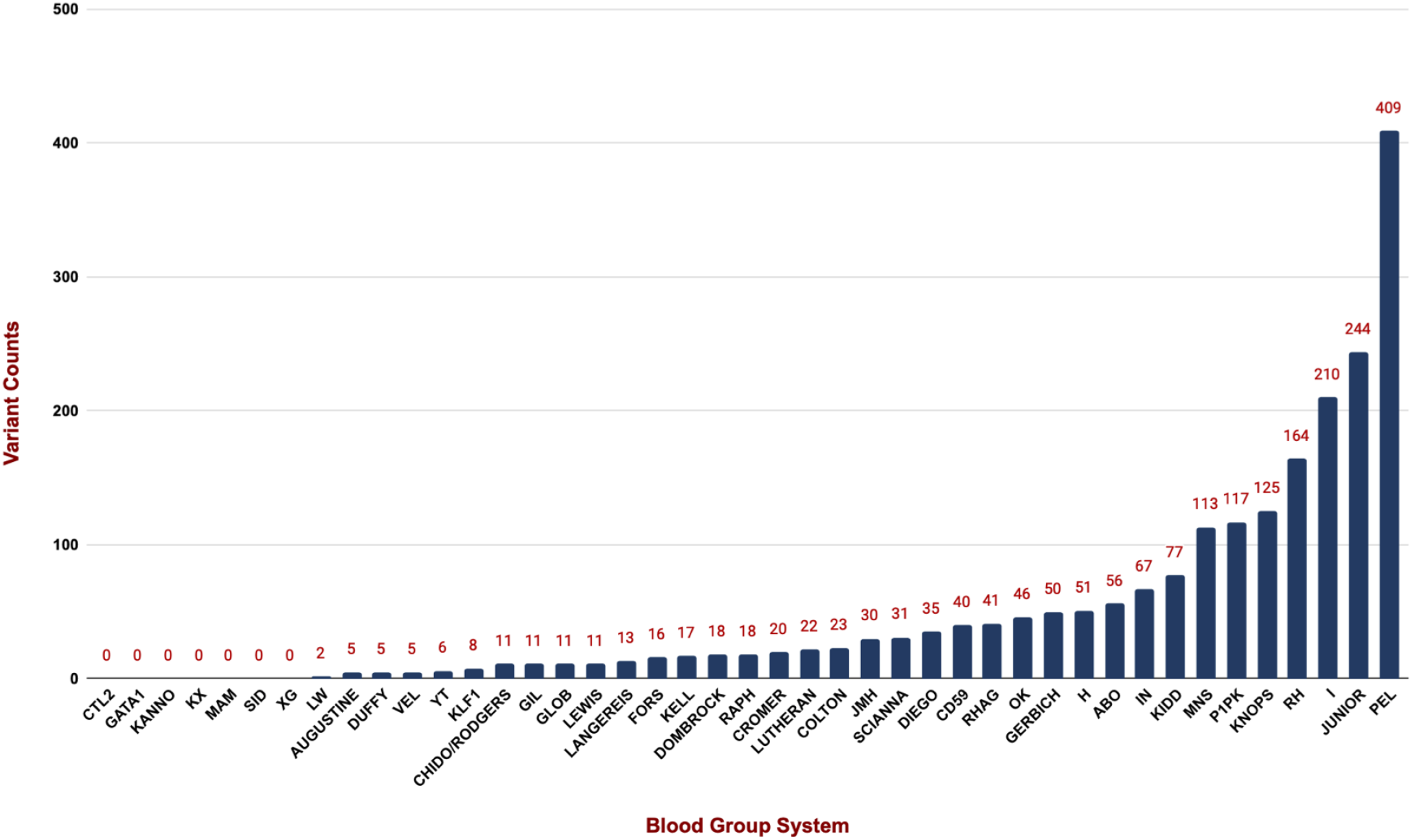
Distribution of potentially novel alleles across the human blood group systems

**Supplementary Table 1**. Brief tabulation of number of variants found associated with human blood group genes in the datasets used in the study. [Summary of blood group variants in study datasets](https://docs.google.com/spreadsheets/d/1imMXrPFsMafdur0RBJgATJWRNKPyAhYZjjq5g9TlfyU/edit#gid=148394494)

**Supplementary Table 2**. Complete blood type profiles of 114 Malaysian Orang Asli samples used in the study [Sample wise complete blood type profiles](https://docs.google.com/spreadsheets/d/1imMXrPFsMafdur0RBJgATJWRNKPyAhYZjjq5g9TlfyU/edit?usp=sharing)

**Supplementary Table 3**. Summary of predicted phenotypes of each blood group system in Malay Orang Asli population [Summary of overall predicted phenotypes of blood group systems](https://docs.google.com/spreadsheets/d/1imMXrPFsMafdur0RBJgATJWRNKPyAhYZjjq5g9TlfyU/edit?usp=sharing)

**Supplementary Table 4**. Distribution of predicted blood phenotypes in Orang Asli subpopulations [Summary of observed blood group phenotypes in various subpopulations of Orang Asli](https://docs.google.com/spreadsheets/d/1imMXrPFsMafdur0RBJgATJWRNKPyAhYZjjq5g9TlfyU/edit#gid=1409681164)

**Supplementary Table 5**. Summary of rare and potentially novel blood group variants in Orang Asli population [List of rare and potentially novel blood group variants](https://docs.google.com/spreadsheets/d/1imMXrPFsMafdur0RBJgATJWRNKPyAhYZjjq5g9TlfyU/edit#gid=657973298)

**Supplementary Table 6**. Frequencies of blood group alleles in various population scale datasets used in this study [Population scale frequencies of filtered blood group variants](https://docs.google.com/spreadsheets/d/1imMXrPFsMafdur0RBJgATJWRNKPyAhYZjjq5g9TlfyU/edit#gid=1356305508)

